# Perception of the consumption of legal and illegal substances in the Honduran population

**DOI:** 10.1101/2023.07.16.23292732

**Authors:** Eleonora Espinoza-Turcios, Carlos Antonio Sosa-Mendoza, Lysien Ivania Zambrano, Henry Noel Castro Ramos, José Armada, Christian R. Mejia

## Abstract

**Introduction:** Substance use affects physical health, mental health, causes social, economic damage in their family environment and society. In recent decades this has changed.

**Aim:** To determine the perception of the consumption of legal and illegal substances and related factors in the general Honduran population.

**Methodology:** Analytical cross-sectional study, through an active search, the information was provided by a suitable informant ≥ 18 years of age, both sexes. Direct questions were asked about consumption, this was associated to influential variables, through descriptive and analytical statistics. Results: Of the 3309 participants distributed nationally, When multivariate analysis was performed, it was found that there was a greater perception of problematic drug use when there was a history of previous violence (aPR: 1.53; 95%CI: 1.22-1.92; p-value<0.001), if cannabis had been consumed in the last quarter (aPR: 1.29; 95%CI: 1.05-1.58; p-value=0.016), if she stopped doing what was expected (aPR : 7.05; 95%CI: 5.00-9.95; p-value<0.001) or if she worried a friend or family member (aPR: 1.49; 95%CI: 1.20-1.86; p-value<0.001), on the contrary, there was less concern among those with university studies (aPR: 0.70; 95%CI: 0.49-0.99; p-value=0.048), adjusted for six variables.

**Discussion:** An association was found between the perception of problematic substance use according to sex, academic grade, history of previous violence, use of tobacco, cannabis, cocaine, opioids, hallucinogens, whether the person stopped doing what was expected, whether a friend or relative was concerned, or whether the person injected.

**Conclusion:** The most reported legal substance of use among the Honduran population was tobacco, followed by marijuana and cocaine among illegal substances.

## Introduction

About 284 million people aged 15-64 years used drugs worldwide in 2020, according to the World Drug Report-2022, an increase of 26% over the previous decade, young people are using more drugs than ever before. In Africa and Latin America, people under 35 years of age account for the majority of those receiving treatment for drug use disorders (UNODC, 2022).Substance use, such as alcohol, tobacco, and illicit drugs, has been associated with serious health problems and is estimated annually to account for 12% of all deaths worldwide (Malaguti et al., 2020).

A 2017 systematic analysis of the global burden of disease shows the largest relative sex differences in 2017 included substance use disorders 3018 cases per 100 000 in men vs 1400 per 100 000 in women, and interpersonal violence 3265 vs 5643, respectively (Sofi-Mahmudi et al., 2021).

According to the World Health Organization (WHO), hazardous use (hazardous drinking) is a pattern of substance use that increases the risk of harmful consequences for the user. Some authors limit these consequences to physical and mental, others also include social consequences (World Health Organization, 1994).

Conditions of poverty, social inequality and violence have exacerbated the problem of substance use as an escape mechanism in Honduras. Although occasional research has been carried out in the past, no recent studies have been conducted in specific communities or in high-risk captive populations. According to a review conducted by the Honduran Institute for the Prevention of Alcoholism, Drug Addiction and Drug Dependency (IHADFA) on drug use in Honduras, it was concluded that the age of onset of substance use has decreased in children, starting at an earlier age. In addition, there is an increase in the prevalence of lifetime use and a significant percentage of young people continue to use. Although illicit drug use is still below the average for the general population, in the homes of these young people there is a high consumption of drugs, mainly alcohol and tobacco, especially by the father figure (“Consumo de drogas en Honduras,” 2001). In view of this situation, we propose to investigate the perception of the consumption of legal and illegal substances and related factors in the general population of Honduras.

## Methods

A cross-sectional analytical or association study was generated, with a non-random convenience sample of 3309 participants, distributed nationally, among the inclusion criteria: informant aged ≥ 18 years, of both sexes, not suffering from any mental illness, giving written informed consent. Exclusion criteria included: suffering from a mental illness that made it impossible to provide information, being intoxicated or having consumed drugs during the interview. In the filtering process, 56 were eliminated because the informant was under 18 years of age, leaving a total of 3309. It should be noted that a statistical power calculation was made to determine whether the number of respondents was sufficient for the crosses made.

An active search for the ideal informant was carried out to try to obtain more reliable information from the consumer, since, due to the social stigma of substance use, the consumer may not give truthful information and may have a bias, the information was obtained through an interview, the recording instrument was anonymous and consisted of: 1) sociodemographic data of the informant. 2) personal background of the consumer. 3) related factors. 4) pathological antecedents of the consumer. The Alcohol, Smoking and Substance Involvement Screening Test (ASSIST), which was developed by the WHO to detect the use of psychoactive substances and related problems in primary care patients, was used to prepare the addiction questions and was validated in a multicenter study conducted in Australia, Brazil, India, Thailand, the United Kingdom, Zimbabwe and the United States (“The Alcohol, Smoking and Substance Involvement Screening Test (ASSIST): manual for use in primary care,” 2010; Humeniuk et al., 2008; WHO ASSIST Working Group, 2002).

The ASSIST, in its version 3.0, is an eight-question instrument of brief application in the last three months that evaluates health risks and other problems associated with substance use (Soto-Brandt et al., 2014).

For the statistical analysis, first a descriptive table was created, where the measures of central tendency and dispersion were generated for the age variable, as well as the frequencies and percentages for the categorical variables. Subsequently, a couple more tables were created and the final tables were generated, where the analytical statistics were performed using generalized linear models (Poisson family, log link function, models for robust variances and with adjustment for the department of residence). In this last part, the PR (prevalence ratios), 95% confidence intervals (95% confidence intervals) and p-values were obtained; it is important to mention that the value p<0.05 was taken as a criterion for a variable to pass from the bivariate to the multivariate model, as well as for statistical significance to be considered in the final model.

The study project was approved by the Biomedical Research Ethics Committee (CEIB) of the FCM IRB 00003070, approved in the session of November 22 of 2018, with registration number: 047-2018, the Doctors in Social Service (MSS) received an online course of Good Clinical Practices The Global Health Network, (www.tghn.org). In addition, it should be noted that at all times the information was safeguarded, the rights of the respondents were respected and followed the ethical guidelines for biomedical research.

## Results

A total of 3309 people participated, distributed in the 18 departments of Honduras, with the highest concentration of participants in the departments of Francisco Morazán 733 (22.2%), Olancho 317 (9.6%) and Comayagua 259 (7.8%). About the people to whom respondents referred, 88.1% (2825) were men, 9.6% (316) were elderly, 55.5% (1796) lived in urban areas, 84.2% (2756) had studies up to high school and 12.5% (410) reported previous violence. Table 1

**Table 1.** Population characteristics, n= 3309.

| Variable | Frequency | Percentage % |
| --- | --- | --- |
| **Gender** |  |  |
| Female | 454 | 13.9 |
| Male | 2825 | 86.1 |
| **Elderly** |  |  |
| No | 2964 | 90.4 |
| Yes | 316 | 9.6 |
| **Resides in** |  |  |
| Rural | 1469 | 45.0 |
| Urban | 1796 | 55.0 |
| **Studies** |  |  |
| Up to high school | 2756 | 84.2 |
| University | 519 | 15.8 |
| **Prior violence** |  |  |
| No | 2872 | 87.5 |
| Yes | 410 | 12.5 |

Regarding the use of legal and illegal drugs in the last quarter, 44.9% (1462) had used tobacco, 8.5% (276) cannabis, 3.6% (116) cocaine, 0.2% (8) opioids and 0.2% (6) hallucinogens. Table 2

**Table 2.** Use of legal and illegal drugs in the last three months, n=3309.

| Variable | Frequency | Percentage % |
| --- | --- | --- |
| **Tobacco\*** |  |  |
| No | 1792 | 55.1 |
| Yes | 1462 | 44.9 |
| **Cannabis\*** |  |  |
| No | 2960 | 91.5 |
| Yes | 276 | 8.5 |
| **Cocaine\*** |  |  |
| No | 3119 | 96.4 |
| Yes | 116 | 3.6 |
| **Opioids\*** |  |  |
| No | 3227 | 99.8 |
| Yes | 8 | 0.2 |
| **Hallucinogens\*** |  |  |
| No | 3229 | 99.8 |
| Yes | 6 | 0.2 |

\*Consumption at least once a month during the last quarter.

In the last quarter 11.7% (388) had some kind of problem derived from drug use, 10.1% (334) stopped doing what was expected, 41.1% (1357) worried a friend or relative, 28.4% (938) tried or succeeded in reducing use, and 0.6% (19) may have injected a drug. Table 3 of your use. **When at least once a month in last trimester stopped doing what was expected. ±Preoccupied a friend or family member in last trimester. £Tried or succeeded in controlling, reducing or stopped using in last trimester. #May have injected a drug in the last trimester.

**Table 3.** Consequences of drug use, n= 3309.

| Variable | Frequency | Percentage % |
| --- | --- | --- |
| **Drug problems\*** |  |  |
| No | 2921 | 88.3 |
| Yes | 388 | 11.7 |
| **Irresponsible\*\*** |  |  |
| No | 2975 | 89.9 |
| Yes | 334 | 10.1 |
| **Concerns‡** |  |  |
| No | 1947 | 58.9 |
| Yes | 1357 | 41.1 |
| **Tried to control£** |  |  |
| No | 2369 | 71.6 |
| Yes | 938 | 28.4 |
| **Injected#** |  |  |
| No | 3286 | 99.4 |
| Yes | 19 | 0.6 |

\* When at least once a month in the last quarter you had some kind of problem because of your use. \*\*When at least once a month in last trimester stopped doing what was expected. ‡Preoccupied a friend or family member in last trimester. £Tried or succeeded in controlling, reducing or stopped using in last trimester. #May have injected a drug in the last trimester.

When the crude analysis was performed, an association was found between the perception of problematic drug use according to sex (p=0.004), academic grade (p<0.001), history of previous violence (p<0.001), whether in the last quarter they had used tobacco (p<0.001), cannabis (p<0.001), cocaine (p<0.001), opioids (p<0.001), hallucinogens (p<0.001), stopped doing what was expected (p<0.001) or worried a friend or relative (p<0.001) or injected (p<0.001). Table 4

**Table 4.**
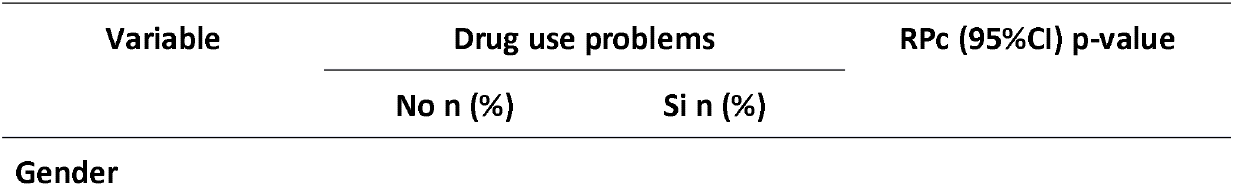
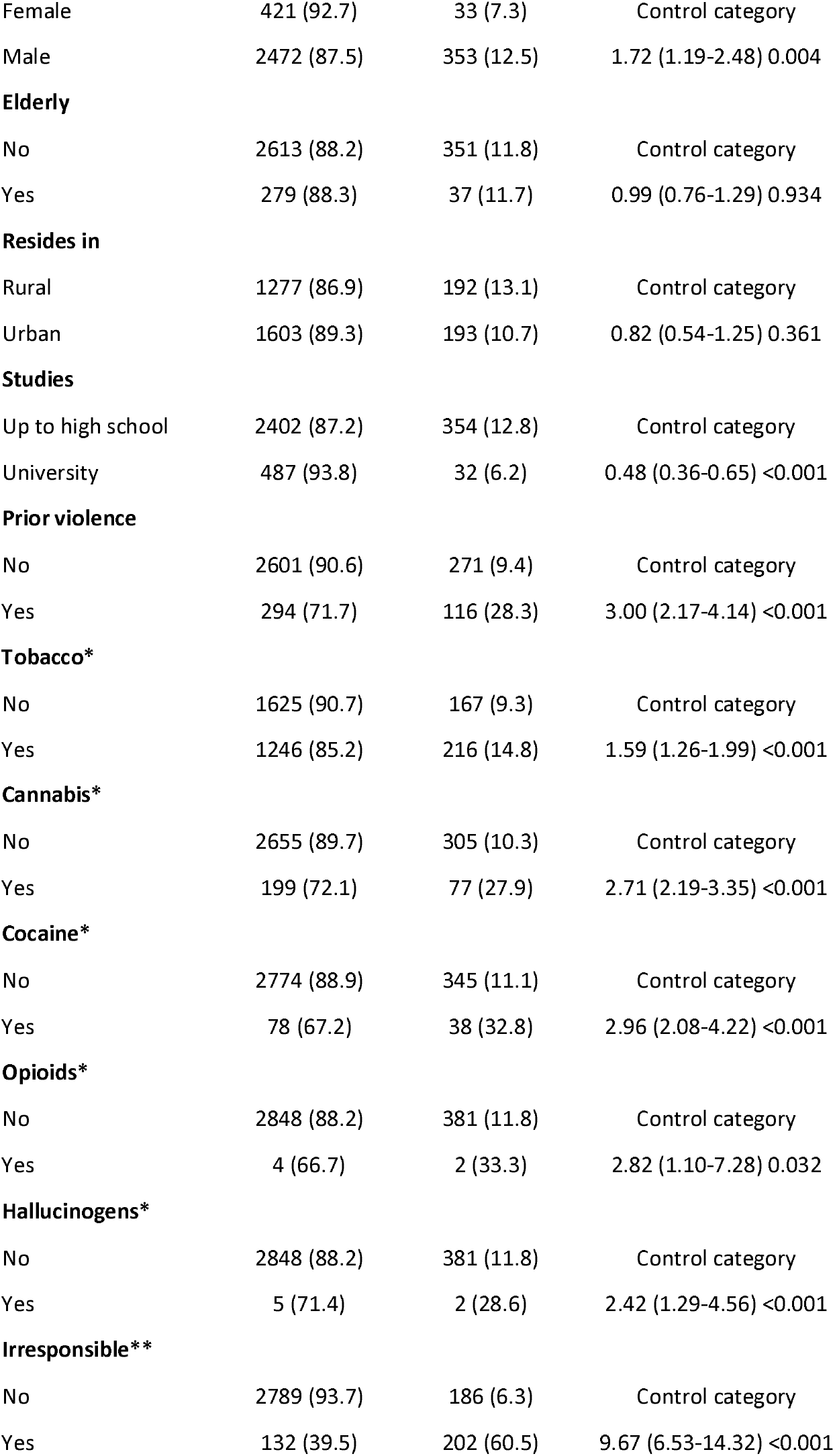
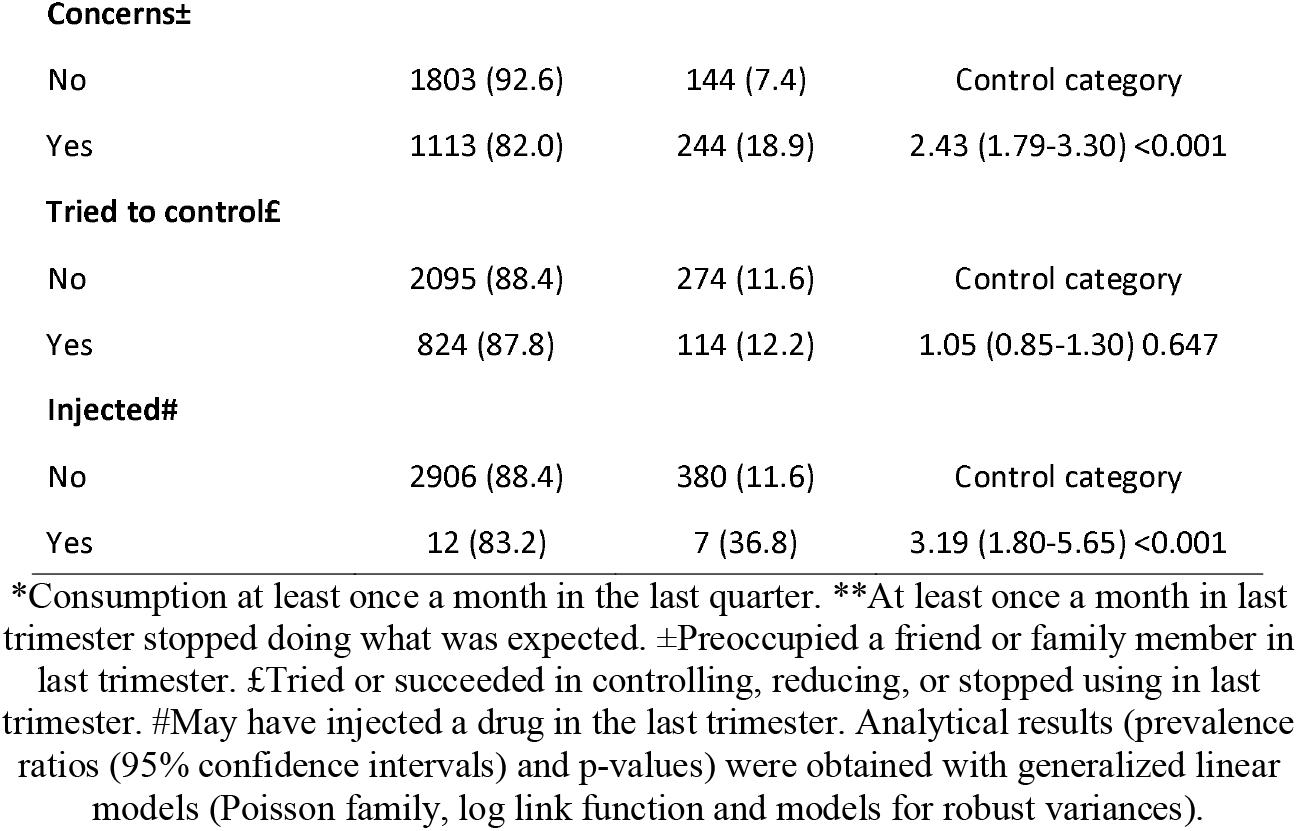
Crude analysis of factors associated with a close relative’s perception of problematic drug use in Honduras, n=3309.

| Variable | Drug use problems |  | RPc (95%CI) p-value |
| --- | --- | --- | --- |
|  | No n (%) | Si n (%) |  |
| Gender |  |  |  |
| Female | 421 (92.7) | 33 (7.3) | Control category |
| Male | 2472 (87.5) | 353 (12.5) | 1.72 (1.19-2.48) 0.004 |
| **Elderly** |  |  |  |
| No | 2613 (88.2) | 351 (11.8) | Control category |
| Yes | 279 (88.3) | 37 (11.7) | 0.99 (0.76-1.29) 0.934 |
| **Resides in** |  |  |  |
| Rural | 1277 (86.9) | 192 (13.1) | Control category |
| Urban | 1603 (89.3) | 193 (10.7) | 0.82 (0.54-1.25) 0.361 |
| **Studies** |  |  |  |
| Up to high school | 2402 (87.2) | 354 (12.8) | Control category |
| University | 487 (93.8) | 32 (6.2) | 0.48 (0.36-0.65) <0.001 |
| **Prior violence** |  |  |  |
| No | 2601 (90.6) | 271 (9.4) | Control category |
| Yes | 294 (71.7) | 116 (28.3) | 3.00 (2.17-4.14) <0.001 |
| **Tobacco\*** |  |  |  |
| No | 1625 (90.7) | 167 (9.3) | Control category |
| Yes | 1246 (85.2) | 216 (14.8) | 1.59 (1.26-1.99) <0.001 |
| **Cannabis\*** |  |  |  |
| No | 2655 (89.7) | 305 (10.3) | Control category |
| Yes | 199 (72.1) | 77 (27.9) | 2.71 (2.19-3.35) <0.001 |
| **Cocaine\*** |  |  |  |
| No | 2774 (88.9) | 345 (11.1) | Control category |
| Yes | 78 (67.2) | 38 (32.8) | 2.96 (2.08-4.22) <0.001 |
| **Opioids\*** |  |  |  |
| No | 2848 (88.2) | 381 (11.8) | Control category |
| Yes | 4 (66.7) | 2 (33.3) | 2.82 (1.10-7.28) 0.032 |
| **Hallucinogens\*** |  |  |  |
| No | 2848 (88.2) | 381 (11.8) | Control category |
| Yes | 5 (71.4) | 2 (28.6) | 2.42 (1.29-4.56) <0.001 |
| **Irresponsible\*\*** |  |  |  |
| No | 2789 (93.7) | 186 (6.3) | Control category |
| Yes | 132 (39.5) | 202 (60.5) | 9.67 (6.53-14.32) <0.001 |
| **Concern±** |  |  |  |
| No | 1803 (92.6) | 144 (7.4) | Control category |
| Yes | 1113 (82.0) | 244 (18.9) | 2.43 (1.79-3.30) <0.001 |
| **Tried to control£** |  |  |  |
| No | 2095 (88.4) | 274 (11.6) | Control category |
| Yes | 824 (87.8) | 114 (12.2) | 1.05 (0.85-1.30) 0.647 |
| **Injected#** |  |  |  |
| No | 2906 (88.4) | 380 (11.6) | Control category |
| Yes | 12 (83.2) | 7 (36.8) | 3.19 (1.80-5.65) <0.001 |

\*Consumption at least once a month in the last quarter. \*\*At least once a month in last trimester stopped doing what was expected. ±Preoccupied a friend or family member in last trimester. £Tried or succeeded in controlling, reducing, or stopped using in last trimester. #May have injected a drug in the last trimester. Analytical results (prevalence ratios (95% confidence intervals) and p-values) were obtained with generalized linear models (Poisson family, log link function and models for robust variances).

Multivariate analysis found that there was a higher perception of problematic drug use when there was a history of previous violence (aPR: 1.53; 95%CI: 1.22-1.92; p-value<0.001), if in the last quarter cannabis use had occurred (aPR: 1.29; 95%CI: 1.05-1.58; p-value=0.016), if it stopped what was expected (aPR: 7.05; 95%CI: 5.00-9.95; p-value<0.001) or if it worried a friend or family member (aPR: 1.49; 95%CI: 1.20-1.86; p-value<0.001), in contrast, there was less worry among those with university education (aPR: 0.70; 95%CI: 0.49-0.99; p-value=0.048), adjusted for six variables. Table 5

**Table 5.**
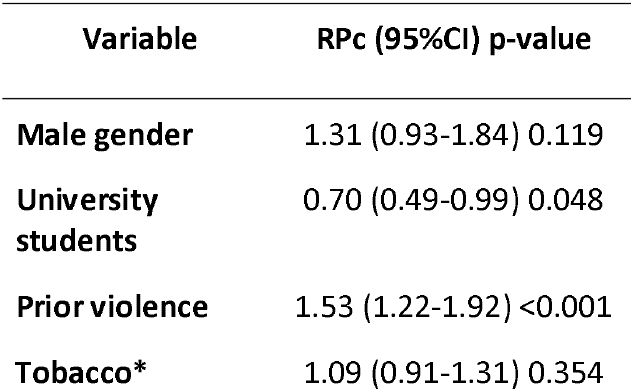
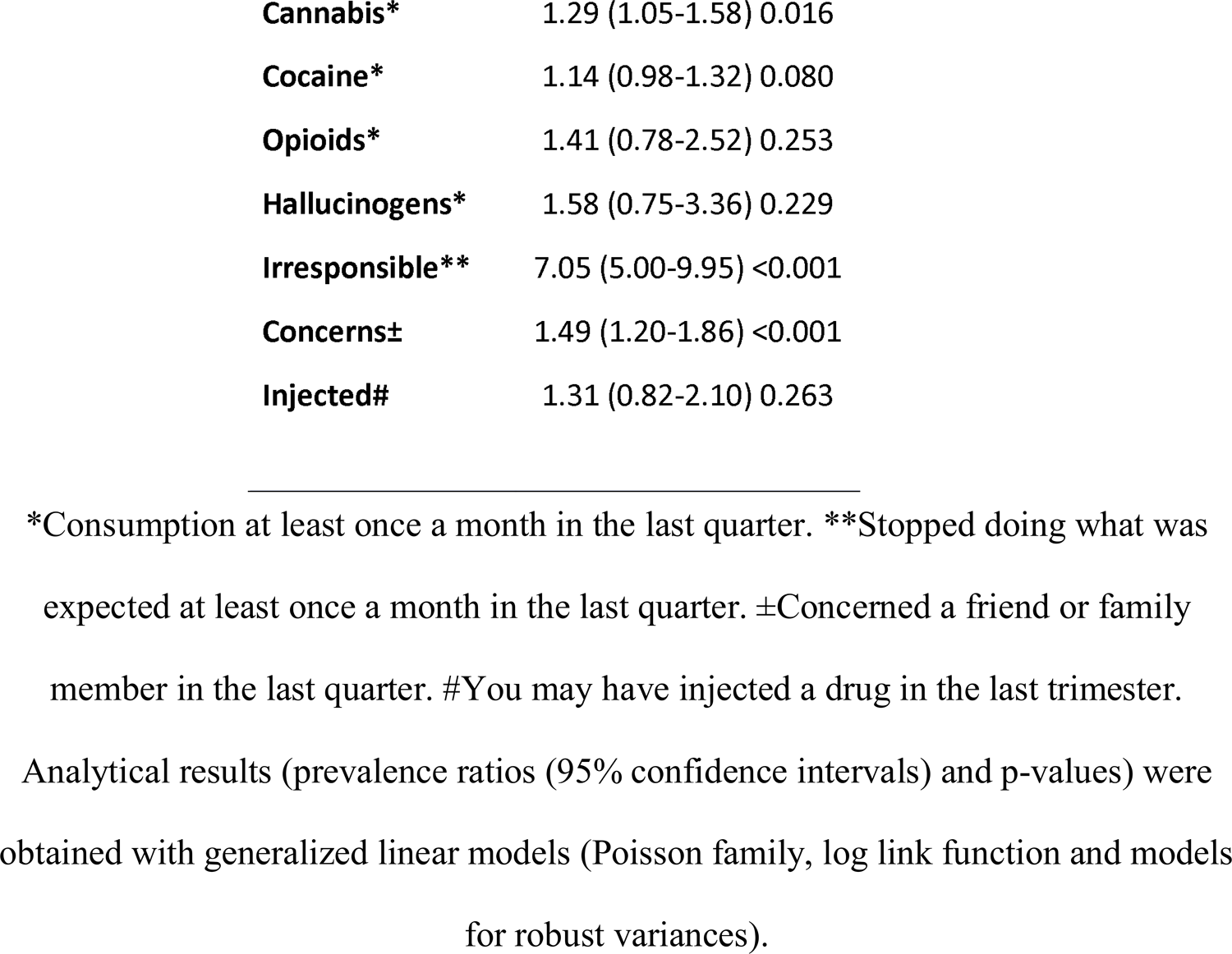
Adjusted analysis of the factors associated with the perception of a close relative of problematic drug use in Honduras.

| Variable | RPc (95%CI) p-value |
| --- | --- |
| Male gender | 1.31 (0.93-1.84) 0.119 |
| University students | 0.70 (0.49-0.99) 0.048 |
| Prior violence | 1.53 (1.22-1.92) <0.001 |
| Tobacco* | 1.09 (0.91-1.31) 0.354 |
| **Cannabis\*** | 1.29 (1.05-1.58) 0.016 |
| **Cocaine\*** | 1.14 (0.98-1.32) 0.080 |
| **Opioids\*** | 1.41 (0.78-2.52) 0.253 |
| **Hallucinogens\*** | 1.58 (0.75-3.36) 0.229 |
| **Irresponsible\*\*** | 7.05 (5.00-9.95) <0.001 |
| **Concerns±** | 1.49 (1.20-1.86) <0.001 |
| **Injected#** | 1.31 (0.82-2.10) 0.263 |

\*Consumption at least once a month in the last quarter. \*\*Stopped doing what was
expected at least once a month in the last quarter. ±Concerned a friend or family
member in the last quarter. #You may have injected a drug in the last trimester.
Analytical results (prevalence ratios (95% confidence intervals) and p-values) were
obtained with generalized linear models (Poisson family, log link function and models
for robust variances).

## Discussion

In recent years in Honduras there has been a considerable increase in the consumption of both legal and illegal substances among the population, the main substances studied being alcohol and tobacco; a study in a locality in the capital of the country showed a prevalence for alcoholism of 6.2% (“Mental disorders prevalence at Villanueva Comunity, metropolitan region,” 1999) According to the National Demographic and Health Survey/Multiple Indicator Cluster Survey (ENDESA/MICS 2019), 31% of men and 9% of women in Honduras consumed alcoholic beverages during the last month. The highest percentages were in the Central District (39%), Bay Islands (38%) and Colon (37%), among those in the 30-34 age group 37%, higher education 42% (INSTITUTO NACIONAL DE ESTADÍSTICA, 2019). In view of this, it was proposed to try to determine the perception of substance use in the Honduran population.

In this study, the highest concentration of participants was registered in the departments of Francisco Morazán 733 (22.2%), Olancho 317 (9.6%) and Comayagua 259 (7.8%); poles of economic development and higher population density, with the most reported substances being tobacco, cannabis, cocaine, opioids and hallucinogens. This is ratified by a study of carriers in the USA in 2022, in the last three months, the substances most used or not used medically were tobacco (82%) for legal drugs, cocaine (53%) for illicit drugs and prescription opioids (67%) for prescription drugs (Tam et al., 2022)

Manthey et al, in an online study with a sample of 36,538 adults from 21 European countries between April and July 2020, found an overall decrease in alcohol use, while cannabis and nicotine use showed increasing trends, as well as cocaine use, but to a lesser extent, while ecstasy use showed a decrease (Manthey et al., 2021). In Spain, the most prevalent drugs of abuse were alcohol, tobacco, hypnosedatives, cannabis and cocaine (“las Toxicomanías (OEDT). Informe 2013: alcohol, tabaco y drogas ilegales en España. Madrid: Ministerio de Sanidad, Servicios Sociales e Igualdad…,” 2015)

A prospective multicenter randomized study with a sample of 3,240 participants reported having used drugs in the last month, 2,084 (64.3%) were identified as having a drug use problem, with the majority of participants reporting most frequent cannabis 59.9%, cocaine 18.1%, street opioids 11.2% or prescription opioids 5.6% (Macias Konstantopoulos et al., 2014).

As reported by Adinolfi et al, 90.86% (587) of the men and 84.55% (1478) of the women had consumed alcoholic beverages, as for illicit drug use, 58.51% (378) of the men and 42.16% (737) of the women had consumed marijuana (Adinolfi, Bezerra, Curado, Souza, & Galduróz, 2022).

In a systematic review the proportion of people who consumed alcohol during the COVID-19 pandemic ranged from 21.7% to 72.9%, this for general population samples, where there was higher use of other substances and ranged from 3.6% to 17.5% in the general population. Mental health factors were the most common correlates or triggers for increased alcohol and other substance use (Roberts et al., 2021).

The abuse of new synthetic opioids is a growing problem worldwide, becoming a serious threat to the health of consumers, these new psychoactive substances can escape immunological screening tests and appropriate confirmatory methods have not yet been developed and validated (Marchei et al., 2018).

In general, it is reported that women drink less than men; however, the absolute number of women who drink has actually increased in the world, 51.4% of women drink alcohol in Europe, while 3.5% of women suffer from alcohol use disorder. (World Health Organization, 2018)

Honduras reports similar patterns in the consumption of substances such as tobacco, cannabis, cocaine; however, in this study we did not find a worrisome increase in alcohol consumption among the population studied; possibly because the information was obtained through a suitable informant.

Hondurans in this study who had prior violence had more drug use, Gilchrist et al. found that 34% of men had been violent in the past year with their current/most recent partner, violence was associated with lower education level, parents who had separated/divorced, childhood physical abuse, alcohol or cocaine use. Intimate partner violence is common among men attending substance abuse treatment (Gilchrist et al., 2015).

General domestic violence and its various types are prevalent in different parts of the world and factors affecting domestic violence, such as general age, age at marriage, low literacy, husband’s addiction to alcohol and drugs, can be prevented through health planning; especially in these areas (Abdi, Mahmoodi, Afsahi, Shaterian, & Rahnemaei, 2021).

Illicit drug use is related to individual characteristics; however, social and family environments seem to be associated with this use (Instituto Brasileiro de Geografia e Estatística-IBGE e o Ministério da Saúde, 2015). Antunes reported that the overall prevalence of illicit drug use was 3.8%; 3.3% among women and 4.4% among men, and physical aggression was by family members and was also associated with illicit drug use (Antunes, Rivadeneira-Guerrero, Goulart, & Oenning, 2018).

Hondurans who used cannabis were also found to have a higher frequency of problematic drug use. Wilkinson et al. reported that marijuana use was significantly associated with worse outcomes on post-traumatic stress disorder (PTSD) symptom severity, violent behavior, and measures of alcohol and drug use (Wilkinson, Stefanovics, & Rosenheck, 2015).

A cross-sectional study using data from the Spanish Household Survey on Alcohol and Drugs (2013) found that men had higher risk consumption of alcohol and cannabis, while women consumed more hypnosedatives. The lower the level of education, the greater the gender differences in the consumption of these substances. While men with a lower level of education had higher risk consumption of alcohol and cannabis, the prevalence in women was the same. Women with a lower level of education and men with a higher level of education consumed more hypnosedatives. Unemployment was associated with substance use in both men and women (Teixidó-Compañó et al., 2018).

In Europe up to 2022, an estimated 83.4 million or 29% of adults (aged 15-64) in the European Union are estimated to have ever used an illicit drug, with more men (50.5 million) than women (33 million) reporting use. Cannabis remains the most widely used substance, with more than 22 million European adults reporting having used cannabis in the past year (“Observatorio Europeo de las Drogas y las Toxicomanías,” 2019).

Pérez and Ruiz 2017, establish that the average age of onset of consumption of psychoactive substances such as cannabis is around nineteen years of age, which is the most prevalent drug after alcohol among the youngest (Pérez & Ruiz, 2017). According to the Colombian Drug Observatory (2016), it was documented that not only is consumption increasing, but the age of onset is decreasing (Restrepo Escobar & Sepúlveda Cardona, 2021).

Among Brazilian adolescents between 13 and 15 years of age, 55.5% reported alcohol consumption and 9.0% had used illicit drugs at least once in their lifetime and 22% reported episodes of drunkenness (Oliveira, Campos, Andreazzi, & Malta, 2017).

In this study according to ASSIST, those who were more irresponsible or worried their friends or family had a higher frequency of drug use, ASSIST screening and linked brief intervention as a project outcome have the potential to reduce the burden of disease associated with substance use and substance use disorders in a wide variety of countries and health care settings (Humeniuk et al., 2012). In Mexico, with a sample of 19,109 Mexicans, with an average age of 34.38 years, men reported high lifetime psychoactive substance use and levels of risky drug use. A high percentage of women aged 18-19 years reported lifetime use of tobacco and alcohol, and a large number of women of all ages reported lifetime use of sedatives and opioids (Morales-Chainé, Robles-García, Barragán-Torres, & Treviño-Santa-Cruz, 2022).

The legalization of cannabis in North America has increased its daily use, especially cannabis products among young adults, in people with psychiatric disorders, suicides and hospitalizations. Cocaine production reached an all-time high in 2020, up 11% over 2019. Methamphetamine trafficking continues to expand geographically; 117 countries reported methamphetamine seizures between 2016 and 2020. Global opium production grew by 7% between 2020 and 2021, reaching 7,930 tons. Illicit drug markets, according to the World Drug Report 2022, can have local, community or individual impacts on the environment (UNODC, 2022)

Mexico has reported that 43.2% of men and 32. 3% of women used drugs in 2021, according to the Mexican Observatory of Mental Health, during the COVID-19 pandemic, 32.5% of the population reported alcohol use, 24.6% tobacco use, and 14.6% cannabis use with higher prevalence in men than in women, 16% of men and 9% of women reported using cocaine, and 16.4% of men and 9.6% of women reported using opioids during the pandemic (Observatorio Mexicano de Salud Mental y Consumo de Drogas, 2021).

A descriptive, cross-sectional, prevalence study to whom the PHQ-9 and the ASSIST 3.0 were applied, in which 50.7% were women with a median age of 41 years. Alcohol was the substance of greatest consumption in the last 3 months with 53.7%, followed by cigarettes 47.6%, marijuana 26.7% and cocaine 14.5% (Campuzano-Cortina et al., 2021). In Rio de Janeiro-Brazil, in another study, prevalences in the last three months were detected in the male subsample for the use of tobacco 56.4%, alcoholic beverages 75.8%, cannabis 16.9% and cocaine/crack 10.1%. Religion and schooling emerge as important factors for drug use (Abreu, Parreira, Souza, & Barroso, 2016).

In contrast, those who were university students were found to have less drug use, with Vanderbruggen et al. reporting that younger individuals with less education or those who were technically unemployed had a higher risk of increasing their alcohol and/or cigarette use during times of social isolation during the COVID-19 pandemic (Vanderbruggen et al., 2020), although other authors report that more schooling increases consumption, as reported in a study in the Norwegian population after the COVID-19 outbreak, where it was reported that the odds of daily alcohol consumption were higher for older people and among those with higher education and lower among women than among men. Daily alcohol consumption was also associated with depression and the expectation of financial loss in relation to the COVID-19 outbreak (Bonsaksen et al., 2021).

Early onset of consumption is directly related to dependence, in our study the onset beginning at early ages from 11 years, having higher risk of dependence in adulthood as being the group of 71-80 years, with 12.0% dependence, according to Pilatti in 2014, reported early onset of alcohol consumption and was associated with the amount of alcohol consumption, even up to dangerous levels, as well as, with tobacco and drug consumption (Pilatti, Caneto, Garimaldi, Del Vera, & Pautassi, 2014).

People with low socioeconomic status (SES) experience a higher risk of mortality in general and alcohol-attributable mortality in particular (Probst, Lange, Kilian, Saul, & Rehm, 2021). Poorer people suffer greater social and health harms from alcohol consumption than wealthier people (World Health Organization -, 2022)

## Limitations

The present study has some limitations that should be taken into account, studying the consumption of both licit and illicit substances due to the social stigma, and therefore, consumers may not provide complete information, which is why we resorted to a suitable informant who may not know the information requested. The concentration of the population was centered in three departments, despite having a sample of all 18 departments, so the results cannot be generalized to the entire population of Honduras, but it could be an important situational analysis.

## Conclusions

Tobacco, followed by cannabis and in third place cocaine were the most commonly used drugs among the Honduran population in the last quarter. Those who had previous violence and those who used cannabis had more problematic drug use and those who were university students had less drug use.

## Author Contributions

Study design: EET, CASM, Data collection: HNCR Data analysis: LIZ, CRM, JA. Writing: EET, CASM, LIZ, HNCR, CRM, JA all authors have read and agreed to the published version of the manuscript.

## Institutional Review Board Statement

The study was conducted under the Declaration of Helsinki. This research’s preparation and execution fully complied with the fundamental ethical principles of autonomy, justice, beneficence, and non-maleficence. The Act Number (2018047), approved by the Ethics Committee in Biomedical Research (CEIB) of the National Autonomous University of Honduras (UNAH), meeting of 22 November 2018.

## Data Availability Statement

The data presented in this study are available on request from the corresponding author.

## Acknowledgments

To all the MSS of the March 2018 to March 2019 cohort who agreed to participate in the project, to the directors of Hospitals and Health Units of the SESAL of the areas that participated in the study, to the teachers of the UIC, for all the support provided in the review of the MSS, assigned to each one. To the memory of Dr. Dagoberto Espinoza Murra (QDG) who was part of the design of the study as an expert in the subject, to Mr. Mauricio Gonzales for the elaboration of the database.

